# Activity of low dose nivolumab in patients with advanced squamous cell carcinomas and other cancers

**DOI:** 10.64898/2026.03.25.26349285

**Authors:** Thibault Gauduchon, Jerome Fayette, Mona Amini-Adle, Eve-Marie Neidhart-Berard, Mehdi Brahmi, Armelle Dufresne, Margaux Dupont, Clelia Coutzac, Axel de Bernardi, Philippe Toussaint, Benoite Mery, Laura Crumbach, Isabelle Ray-Coquard, Aurélie Dutour, Marie Castets, Jean-Yves Blay, Pierre Etienne Heudel

## Abstract

Immune checkpoint inhibitors such as anti–PD-1 antibodies are essential in cancer therapy. Emerging data suggest that lower doses may be effective and more economical, though further evidence is needed. We conducted a retrospective study at Centre Léon Bérard to assess the efficacy and safety of low-dose nivolumab (20 mg every three weeks) in patients with advanced cancer, mainly squamous cell carcinomas (SCC). Between 2023 and 2024, 53 patients were treated, with a median age of 74 years; 39.6% were over 80. Most were male (64%) and had ECOG ≥2 (69.9%). Primary tumor sites included cutaneous SCC (34%), head and neck SCC (32%), and soft tissue sarcoma (15%). After a median follow-up of 8.3 months, median overall survival was 7.5 months. The objective response rate (ORR) was 20.8% overall, rising to 35.3% in cutaneous SCC and 23.5% in head and neck SCC—comparable to standard-dose nivolumab. Toxicity was manageable: 18.7% experienced immune-related adverse events, with only 3.7% grade 3. Low-dose nivolumab demonstrates encouraging efficacy and tolerability in a frail population, supporting its potential role in resource-limited settings. Prospective trials are warranted to confirm these findings in broader populations.

## INTRODUCTION

Within the last 15 years, immunotherapy with immune checkpoint blockers have emerged as a cornerstone in the treatment of a large and still growing number of malignancies (1,2). These are now used in first line setting in many advanced neoplastic diseases, as well as in adjuvant and neoadjuvant setting eg in melanoma, NSCLC, or MSI-H colorectal cancers (3-10). Among others, nivolumab, a programmed death-1 (PD-1) inhibitor, has demonstrated efficacy in a broad spectrum of neoplastic diseases. The recommended dose for nivolumab is 3mg/kg or a flat dose of 240mg/ 480 mg every 2 to 4 week course (3-9). However, lower doses of nivolumab from 0.1 to 1 mg/kg are biologically capable to block the PD-1 signalling and to be clinically active (11-15). PD-1 receptor occupancy of 70% was reported to be achieved by nivolumab at a dose low as 0.3 mg/kg. The clinical efficacy and side effect profile of nivolumab was reported to be comparable at lower doses (14,15).

Several studies have shown that reduced-dose nivolumab can be effective across different cancers. In NSCLC, low doses (20–100 mg every three weeks) achieved similar objective response rates and survival compared to standard dosing [16]. Patil et al. found that 20 mg nivolumab combined with metronomic chemotherapy significantly improved one-year overall survival in advanced HNSCC (from 16% to 43%) without added toxicity [17]. In classical Hodgkin lymphoma, a phase II trial using low-dose nivolumab with AVD as first-line therapy reported a 100% objective response rate and 65% complete remissions, while reducing treatment costs [18]. Another study at ESMO Asia 2024 showed improved progression-free survival in metastatic solid tumors with early low-dose nivolumab (40 mg), with two-thirds of patients experiencing no side effects [19]. Pharmacokinetic analyses confirmed similar half-life and drug exposure between low-dose and standard-dose regimens, with PD-1 receptor saturation occurring even at very low doses [20]. Plasma levels of nivolumab remained above the therapeutic threshold in both dosing groups. Clinical outcomes such as progression-free survival and response rates were comparable, while grade ≥3 adverse events were lower with low doses (0% vs. 28.57%). These findings suggest that low-dose nivolumab maintains efficacy with better safety. Ongoing trials, including a phase II study in India, are now evaluating low-dose nivolumab (40 mg) with chemotherapy as neoadjuvant treatment for resectable NSCLC [21].

The safety profile of low dose Nivolumab seems to be better and represent an opportunity to treat fragile, old patients with comorbidities. Several retrospective, prospective, and randomized controlled studies have thus investigated 10-fold lower doses considering the overall cost of this treatment for health care systems, applied to rapidly growing numbers of malignancies (16-28). These studies showed a significant antitumor activity in a variety of cancers. This approach could be particularly beneficial for patients who cannot have access to standard dosing regimen due to financial constraints, but also existing comorbidities (11, 14,15). When anti-PD1 is not accessible for cost issues, when signals of activity are reported in disease where the anti-PD1 is not formally reimbursed, the use of low dose PD1Ab may represent a therapeutic option (27).

Despite encouraging preliminary data, evidence remains scarce regarding the outcomes of low-dose nivolumab across different tumor types. We present a comprehensive retrospective series of 53 patients treated with low-dose nivolumab at our center since February 2023. Treatment was proposed by multidisciplinary tumor boards for patients who had exhausted standard therapeutic options or for whom nivolumab was not reimbursed despite documented efficacy at conventional doses. This report outlines the clinical characteristics and outcomes of these patients, highlighting those who derived benefit from this alternative strategy.

## PATIENTS AND METHODS

### Patients

All patients treated with low-dose nivolumab at the Centre Léon Bérard (CLB) since 2023 were included in a retrospective study. Inclusion criteria were patients treated with low-dose nivolumab as a specific line of treatment, excluding those in whom nivolumab dose reduction was implemented due to toxicity. Patients previously treated with immune checkpoint inhibitors (ICIs) at standard doses—either alone or in combination with chemotherapy or radiotherapy—were included in the analysis only if they had received at least one systemic treatment line without ICIs before initiating low-dose nivolumab as a rechallenge.

### Clinical and biological data

The data were collected using the CONSORE tool from the electronic patient records of the CLB (29-31). Fifty three patients were treated with low dose nivolumab 20mg course every 21 days in the CLB from Feb 2023. Patient characteristics, treatments and biological results were extracted and checked manually by 3 of us (TG, JYB and PH). Clinical characteristics (age, gender, histological type, PS, weight, lines of treatment…) and biological (PD-L1 expression, tumor mutational burden, MSI-H status, when available) characteristics of all patients were collected at baseline, extracted, verified, and validated. Survival, response to treatment according to RECIST, time to progression or next treatment or death, and follow-up were collected.

### Nivolumab administration

Nivolumab was administered IV at the dose of 20mg flat dose in in 100 mL normal saline over 60 minutes once every 3 weeks.

### Ethical approval and health authorities approval

This retrospective study was conducted in accordance with the MR004 methodology of the French National Health Authorities and was approved on December 23, 2024 (R201-004-521) receiving validation from the local data protection officer, on behalf of French regulatory authorities. All patients were informed of the potential use of their health data for research purposes and did not express opposition.

### Statistical analysis

The distribution of clinical and biological characteristics was analyzed using the chi square test, Fisher exact test, Mann-Whitney U test. Survival was plotted from the date of nivolumab injection to the date of death or to the date of last news if alive at the time of the analysis (March 2025). Survival was plotted according to the Kaplan-Meier method, and groups were compared using the logrank test. All statistical analyses will be performed using SPSS 23.0 software SPSS (IBM, Paris, France).

## RESULTS

### Description of patients

The study cohort consisted of 53 patients, all diagnosed with metastatic disease. The median age was 74 years (range: 20-97 years). Twenty-one patients (39.6%) were over 80 years old. The majority were male (34 patients, 64%). The most common primary cancer sites included squamous cell carcinomas (SCC) (18 patients, 34%), head and neck cancers (17 patients, 32%), sarcomas (8 patients, 15%), colorectal cancer (3 patients, 5.7%), and vulvar cancer (2 patients, 3.8%). Performance status (PS) was assessed at baseline, on day 1 of the first nivolumab injection. The majority of patients (37 patients, 69.8%) had an ECOG performance status (PS) of ≥2, indicating significant functional impairment. Among them, 32 patients (60.4%) had a PS of 2, 4 patients (7.5%) had a PS of 3, and 1 patient (1.9%) had a PS of 4. Regarding prior lines of therapy, 29 patients (54.7%) received nivolumab as a third-line or later treatment. Additionally, 16 patients (30.1%) had previously been treated with standard-dose immune checkpoint inhibitors, reflecting a pre-exposed population with potentially lower responsiveness to further immunotherapy. Nivolumab was administered at a low fixed dose of 20 mg every three weeks, with patients receiving a median of three doses (range: 1–18 doses). Notably, 12 patients (22.6%) received only one course of low-dose nivolumab, indicating early discontinuation due to continuous disease progression or worsening of the patient’s general condition. At the time of this analysis, only 10 patients were able to receive an additional line of treatment after low-dose nivolumab.

### Response rates, survival and toxicity

With a median follow-up of 8.3 months, the median overall survival (OS) was 7.5 months in the series (Figure 1A, Table 1). Performance status >1 was the only parameter correlated to survival (Table 1). Figure 1A-D shows the OS and the time to progression, subsequent treatment, or death for all patients (Fig 1A-B), as well as in the squamous cell carcinoma group and the non-squamous cell group (Figure 1C-D).

**Table 1:**
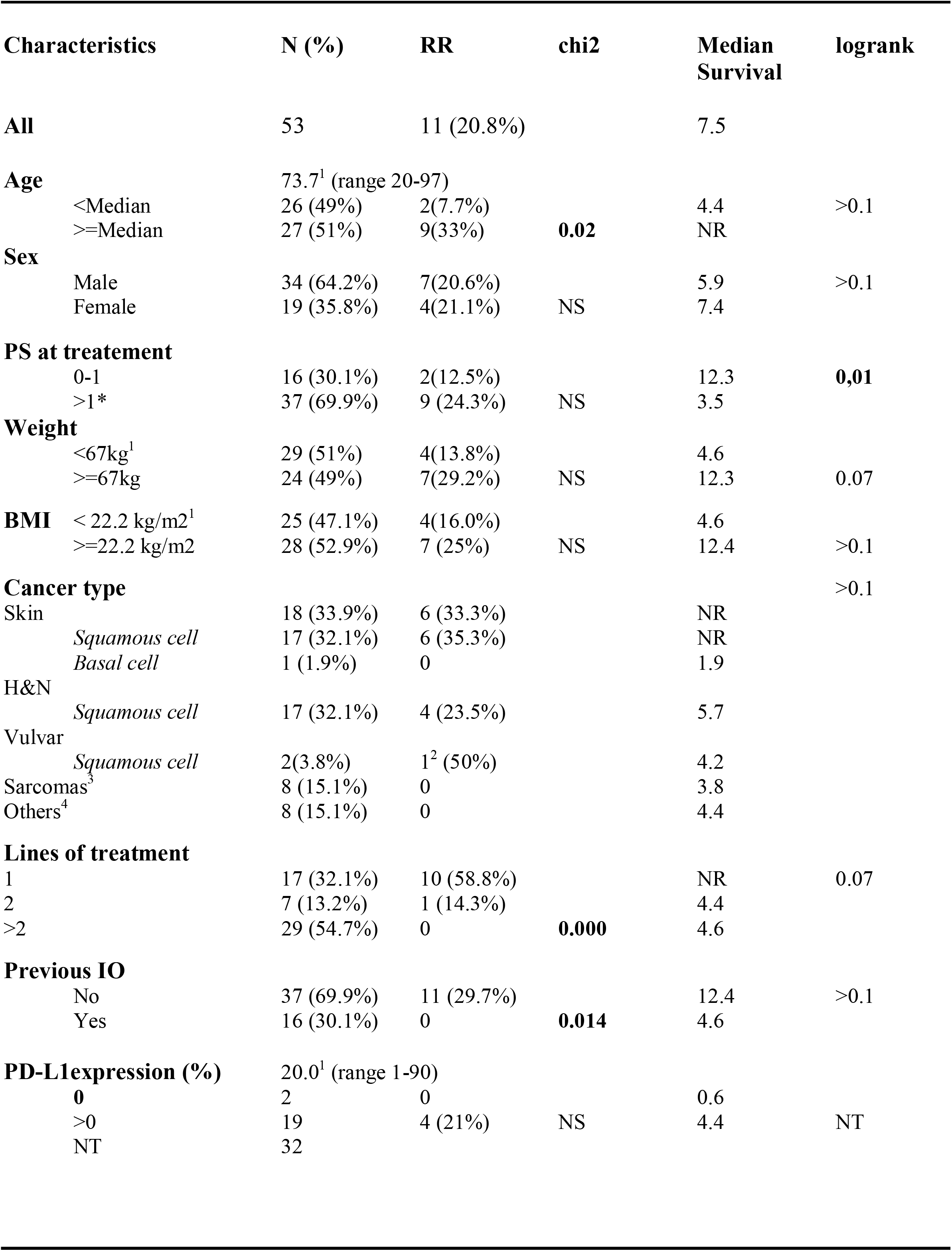
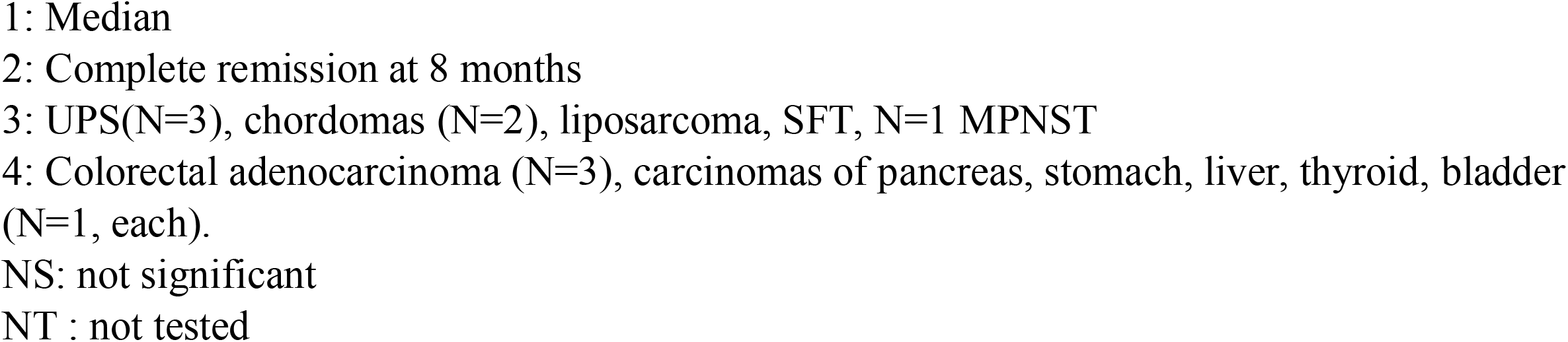
Characteristics of the patients.

**Figure 1:**
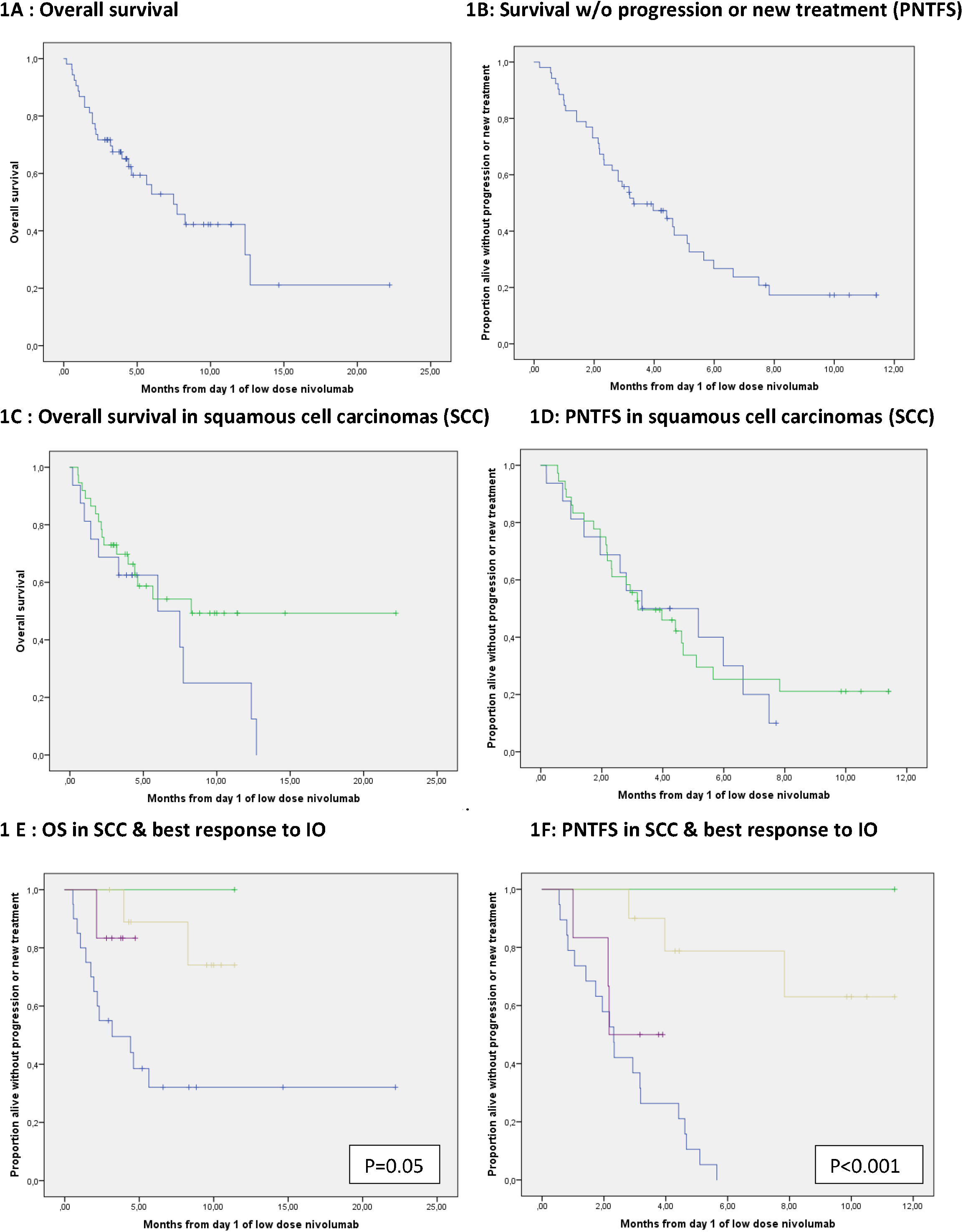
Survival and time to next treatment in the different subgroups. **Legend figure 1** 1A: Overall survival from the start of low dose nivolumab 1B: Survival without progression or new treatment (PNTFS) after the start of low dose nivolumab 1C: Overall survival in squamous cell carcinoma (skin, head and neck or vulva in green) and other cancer histotypes(in blue) 1D: Survival without progression or new treatment from the start of low dose nivolumab in squamous cell carcinoma (skin, head and neck or vulva, green) and other cancer types (blue) 1E: Overall survival in squamous cell carcinoma (skin, head and neck or vulva) according to response: PR (brown) or CR (green), Stable disease (SD) (purple) or progressive disease (blue) 1F: Survival without progression or new treatment after the start of low dose nivolumab in squamous cell carcinoma (skin, head and neck or vulva) according to response to low dose nivolumab: PR (brown) or CR (green), stable disease (SD) (purple) or progressive disease (blue) Proportion alive without progression or new treatment Months from day 1 of low dose nivolumab

The response rate was 11 out of 53 (20.8%), including 3 complete responses (CR) out of 53 (5,6%), observed in 2 cutaneous squamous cell carcinoma (cSCC) and in a case of vulvar carcinoma in the second-line setting. The median duration of response was 7.8 months (range: 2.8–11.4). All responses were observed in patients with squamous cell carcinoma (N=37) including 17 cSCC (ORR= 6/17, 35.3%); head and neck SCC (ORR =23.5%, 4/17) and vulvar SCC (1/2; 50%)). For this subgroup of 37 patients, the RR was 11/37 (29.7%) vs 0/16 for the other patients (p=0.014). With a median follow-up of 8.5 months, 3 of the 11 patients with squamous cell carcinomas in complete or partial response have ultimately progressed, and two died from progressive disease (Figure 1E-F). No objective responses were observed in the sarcoma subgroup. One of the skin squamous cell carcinoma patient of this series achieved tumor control (SD) with standard dose ICI after progressing under low dose.

Responses in this subgroup were observed in patients with PS 0 to 2, in first or second line of treatment only, but in none of the patients with previous IO (Table 1). The subgroup of 22 patients with squamous cell carcinoma in first or second line, naive of IO, had a response rate of 11/22 (50%) with 3 (13.6%) stable disease as best responses.

Among the 21 tested patients, PD-L1 expression was ≥1% in 19 (90.5%) on primary tumor cells. Response rate was 4/19 (21.1%) in patients with detectable PD-L1 vs 0/2 in patients with PD-L1 negative cancers. TMB was available in 4 patients (<1,<1, 6, 9.6 mut/megabase respectively): none were responders. A single patient had a MSI-H colorectal cancer (among the 5 tested in the series); previously treated with pembrolizumab, he had PD as best response to low dose nivolumab.

Among 53 patients, 10 experienced immune-related adverse events (IRaes) associated with low-dose nivolumab treatment. Two patients (3.7%) presented grade 3 toxicity including one transient increased transminases spontaneously regressive without permanent IO discontinuation and one myocarditis. Eight other patients (15%) presented with grade 1 toxicity including arthralgia (n=3), skin rash (n=2), diarrhea (N=2) and myositis (n=1) One presented a stroke considered unrelated. Two patients were diagnosed with a different cancer after respectively 1 and 2 injections of low dose nivolumab (one stage 4 colon carcinoma, one skin melanoma), both considered unrelated to low dose nivolumab by the treating physician.

### Subgroup Analysis of Cutaneous Squamous Cell Carcinoma

We conducted a subgroup analysis of patients with recurrent/unresectable cSCC who received at least 2 cycles of low-dose nivolumab (20 mg q3w) between 08/12/2023 and 17/01/2025. A total of 12 patients (lymphadenopathy: n=4, periauricular: n=3, temporal: n=2, vertex: n=1, other: n=2) with a mean age of 86.6 years received an average of 6.6 cycles of low-dose nivolumab. The 6-month PFS rate was 73% (90% CI: 0.51– 1.00). Among the 10 evaluable patients, 7 achieved a response (2 complete and 5 partial), resulting in an overall response rate of 58% (90% CI: 31–81%). 3 patients experienced disease progression, while 2 were not yet evaluable (Figure 2). The treatment was well tolerated, with no severe adverse events or treatment discontinuations due to toxicity.

**Figure 2:**
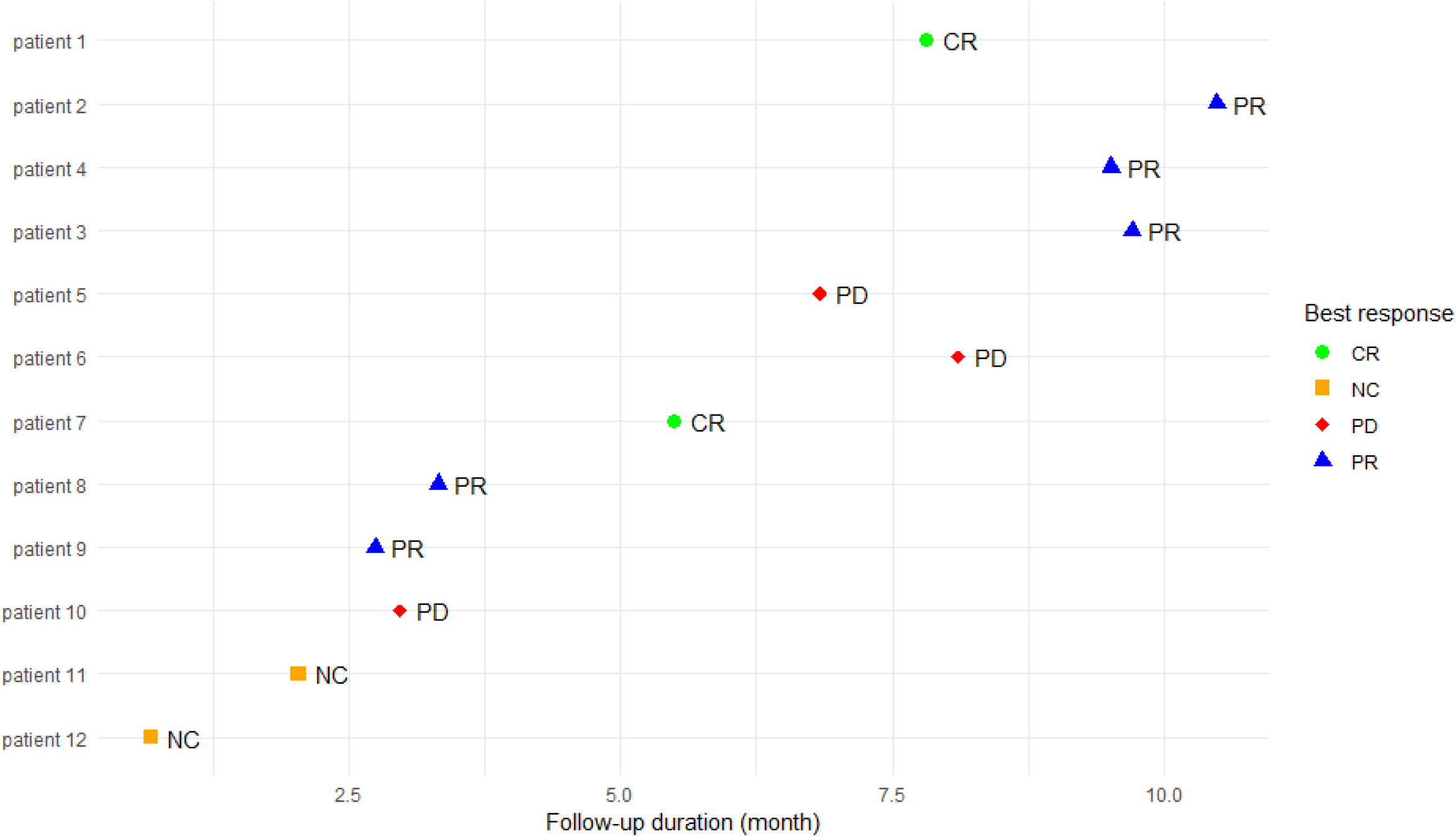
Follow-up of patients with Cutaneous Squamous Cell Carcinoma. **Legend Figure 2** CR: Complete Response PR: Partial Response PD: Progressive Disease UR: Unknown Response

## DISCUSSION

This study reports our experience with single-agent low-dose immune checkpoint inhibition using nivolumab at 20□mg every three weeks in metastatic cancer patients who had no access to standard-dose immunotherapy (IO). Conducted as a monocentric, retrospective analysis, the study included all patients treated with low-dose nivolumab during the inclusion period. Treatment selection was based on previously published evidence supporting the use of low-dose nivolumab, and it was offered to patients with cancers for which standard-dose ICI therapy was not reimbursed at the time of prescription [11-27]. Given prior reports of a favorable safety profile at lower doses, this strategy aimed to provide a feasible and accessible therapeutic alternative. The overall response rate (ORR) of 20.8% (11/53) in the entire cohort—and notably 29.7% among patients with squamous cell carcinoma (SCC)—suggests that low-dose nivolumab retains meaningful antitumor activity, particularly in cutaneous and head and neck SCC. All responses were observed in SCC patients, with an ORR of 50% in those treated in the first or second line, and without prior exposure to ICIs. These findings are consistent with earlier studies indicating that PD-1 receptor occupancy—and thus therapeutic efficacy—can be achieved even at low serum concentrations of nivolumab.

### Efficacy in a Frail Population

The majority of patients in this study represented a frail population, with a median age of 73 years, a performance status (PS) of 2 or higher, and a history of multiple prior treatments. Most would not have qualified for clinical trials due to age, PS, or comorbidities. As expected, the median overall survival was limited at 7.5 months. However, a subset of long-term responders (>15 months) emerged among SCC patients, supporting the potential of low-dose nivolumab in this subgroup. Among the 22 patients with metastatic SCC treated in the first or second line, without prior IO exposure, 11 (50%) achieved an objective response. These responses occurred regardless of age or PS; however, all responders had a PS between 0 and 2. Given the combination of observed activity and tolerability in this subgroup, prospective studies are warranted to further explore low-dose nivolumab, ideally with inclusive eligibility criteria accounting for age and performance status.

### Safety and Tolerability

Toxicity, as recorded in the electronic health records, was limited. Two patients experienced grade 3 adverse events (elevated transaminases and cardiotoxicity) leading to treatment interruption, and both were considered related to nivolumab. While grade 3 toxicities appeared less frequent than those typically seen with standard dosing, they were not absent, in line with previous reports. Other toxicities were mild (grade 1), and treatment discontinuation occurred predominantly due to disease progression rather than toxicity. Nonetheless, the retrospective nature of the study may have led to underreporting of adverse events, and future prospective evaluations will be important to better define the safety profile.

### Clinical and Economic Implications

Beyond clinical efficacy, the use of low-dose nivolumab carries significant economic relevance. As the cost of immunotherapy continues to rise globally, particularly in low- and middle-income countries, dose optimization emerges as a compelling strategy to enhance access while preserving therapeutic benefit. Several pharmacokinetic studies have demonstrated that lower doses of nivolumab achieve comparable PD-1 receptor saturation and similar exposure metrics to standard doses (11–13), reinforcing the rationale for personalized, cost-effective dosing strategies. In healthcare settings where nivolumab is not reimbursed for specific indications, low-dose regimens may offer a pragmatic and affordable alternative. Our findings suggest that, even in the absence of reimbursement, patients with squamous cell carcinoma—especially when treated in earlier lines—may still derive clinically meaningful benefit from low-dose immunotherapy. This approach could help bridge treatment gaps and reduce disparities in access to innovative cancer therapies.

### Limitations and Future Directions

This study has several limitations inherent to its retrospective, single-center design. The cohort included patients who were potential candidates for immunotherapy but were ineligible for standard treatment due to reimbursement constraints. The population was clinically heterogeneous, including various tumor types, performance status levels, and prior lines of therapy, which may confound interpretation of outcomes. Toxicity data were collected retrospectively and may underestimate the true incidence of adverse events. Additionally, the impact of treatment on quality of life could not be assessed due to the study design. Although the observed activity in squamous cell carcinoma—particularly in earlier treatment lines—was notable, these findings require prospective validation. It remains unclear whether the efficacy observed is exclusive to SCC histology or if other tumor types may also benefit from low-dose strategies. Dedicated prospective trials across various histologies are warranted to explore this further. Importantly, the current study does not provide a direct comparison between standard and low-dose nivolumab in terms of efficacy or safety. Such comparisons should be addressed in future randomized studies, ideally incorporating a crossover design for patients progressing on low-dose therapy. Despite these limitations, our findings contribute to the growing body of evidence supporting the feasibility of low-dose nivolumab in a frail, often undertreated population. Most responders had a PS of 2 and a median age of 84 years, emphasizing the relevance of this approach in elderly or comorbid patients. Notably, no responses were observed beyond the second line of therapy, highlighting the importance of early use in eligible patients. Finally, the potential cost savings associated with this strategy merit further investigation, particularly in healthcare systems facing economic constraints.

## CONCLUSION

In conclusion, these results show that low dose nivolumab yields tumor responses in adult patients with advanced cancer, particularly those with squamous cell carcinomas, in an elderly and mostly frail population. The toxicity profile was excellent. Prospective evaluation of this dose vs standard dose of IO deserve to be further investigated in a prospective randomized clinical trial, ideally with a cross over design.

## Data Availability

All data produced in the present study are available upon reasonable request to the corresponding author. Data include anonymized patient records from the Centre Leon Berard.

## REFERENCES

1. Pardoll, D.M. The blockade of immune checkpoints in cancerimmunotherapy. Nat. Rev. Cancer 12, 252–264 (2012).

2. Chen, D.S. & Mellman, I. Oncology meets immunology: the cancer-immunity cycle. Immunity 39, 1–10 (2013).

3. Weber J, Mandala M, Del Vecchio M, Gogas HJ, Arance AM, Cowey CL, et al s. Adjuvant Nivolumab versus Ipilimumab in Resected Stage III or IV Melanoma. N Engl J Med. 2017; 377:1824–1835.

4. Brahmer J, eckamp KL, Baas P, Crinò L, Eberhardt WE, Poddubskaya E,et al. Nivolumab versus docetaxel in advanced squamous-cell non-small-cell lung cancer. N. Engl. J. Med. 2015; 373, 123–135.

5. Borghaei H, et al. Nivolumab versus docetaxel in advancednonsquamous non-small-cell lung cancer. N. Engl. J. Med. 2015 ; 373,1627–1639.

6. Motzer, R.J. Escudier B, McDermott DF, George S, Hammers HJ, Srinivas S,et al. Nivolumab versus everolimus in advanced renal-cell carcinoma. N. Engl. J. Med. 373, 1803–1813.

7. Ansell SM. Nivolumab in the Treatment of Hodgkin Lymphoma. Clin Cancer Res. 2017; 23:1623–1626.).

8. Patel SP, Othus M, Chen Y, Wright GP Jr, Yost KJ, Hyngstrom JR, et al. Neoadjuvant-Adjuvant or Adjuvant-Only Pembrolizumab in Advanced Melanoma. N Engl J Med. 2023 Mar 2;388(9):813–823.

9. Blank CU, Lucas MW, Scolyer RA, van de Wiel BA, Menzies AM, Lopez-Yurda M,et al. Neoadjuvant Nivolumab and Ipilimumab in Resectable Stage III Melanoma. N Engl J Med. 2024; 391:1696–1708.

10. Le DT, Uram JN, Wang H, et al: PD-1 blockade in Tumors with mismatch-repair deficiency. N Engl J Med 2015 ; 372:2509–2520.

11. Brahmer JR, Drake CG, Wollner I, et al: Phase I study of single-agent anti–programmed death-1 (MDX-1106) in refractory solid tumors: Safety, clinical activity, pharmacodynamics, and immunologic correlates. J Clin Oncol 28:3167–3175, 2010

12. Agrawal S, Feng Y, Roy A, et al: Nivolumab dose selection: Challenges, opportunities, and lessons learned for cancer immunotherapy. J Immunother Cancer 4:72, 2016

13. Sheng J Srivastava S, Sanghavi K, et al: Clinical pharmacology considerations for the development of immune checkpoint inhibitors. J Clin Pharmacol 57:S26–S42, 2017 (suppl 10)

14. Feng Y, Wang X, Bajaj G, et al: Nivolumab exposure–response analyses of efficacy and safety in previously treated squamous or nonsquamous non–small cell lung cancer. Clin Cancer Res 23: 5394–5405, 2017

15. Prabhash, K.; Patil, V.M.; Noronha, V.; Joshi, A.; Abhyankar, A.; Menon, N.; Banavali, S.; Gupta, S. Low doses in immunotherapy: Are they effective? Cancer Res. Stat. Treat. 2019, 2: 54.

16. Yoo, S.H.; Keam, B.; Kim, M.; Kim, S.H.; Kim, Y.J.; Kim, T.M.; Kim, D.-W.; Lee, J.S.; Heo, D.S. Low-dose nivolumab can be effective in non-small cell lung cancer: Alternative option for financial toxicity. ESMO Open 2018, 3, e000332.

17. Patil VM, Noronha V, Menon N, et al: Low-dose immunotherapy in Head and neck cancer: A randomized study. J Clin Oncol 2023; 41:222–232.

18. Vega Mateos A, Vaquera Alfaro HA, Tejada Vasquez AC, Lopez-Garcia YK, Garcia-Salas G, Cantu O, et al. Low-Dose Nivolumab Plus AVD As Front-Line Therapy for Classical Hodgkin’s Lymphoma: Preliminary Results of a Phase 2 Trial. Blood. 5 nov 2024;144(Supplement 1):1665.

19. Melemadathil, K.R. et al. 472P Exploring the efficacy of low-dose nivolumab in metastatic settings. Annals of Oncology, Volume 35, S1577

20. Gandhi KA, Shirsat A, Hj SK, Chavan A, Dicholkar P, Shah S, et al. Pharmacokinetics and clinical outcomes of low-dose nivolumab relative to conventional dose in patients with advanced cancer. Cancer Chemother Pharmacol. nov 2024;94(5):6591Z68.

21. Biswal N. 613TiP Evaluation of low dose Nivolumab with neoadjuvant chemotherapy in non-small cell lung cancer: A phase II open label randomised clinical trial (ELON). Ann Oncol. 1 déc 2024;35:S1622.

22. Prabhash, K.; Abraham, G.; Menon, N.; Patil, V.M.; Joshi, A.P. The efficacy of low-dose immunotherapy in head-and-neck cancer. Cancer Res. Stat. Treat. 2019, 2, 268.

23. Zhao JJ, Kumarakulasinghe NB, Muthu V, et al: Low-dose nivolumab in renal cell carcinoma: A real-world experience. Oncology 99:192–202, 2021 19.

24. Kumar, A.; Noronha, V.; Patil, V.; Joshi, A.; Menon, N.; Kapoor, A.; Janu, A.; Mahajan, A.; Rajendra, A.; Prabhash, K. 1049P Efficacy and safety of low dose immunotherapy in palliative setting of advanced solid tumours. Ann. Oncol. 2020, 31, S718.

25. Zhao, J.J.; Kumarakulasinghe, N.B.; Muthu, V.; Lee, M.; Walsh, R.; Low, J.L.; Choo, J.; Tan, H.L.; Chong, W.Q.; Ang, Y.; et al. Low-Dose Nivolumab in Renal Cell Carcinoma: A Real-World Experience. Oncology 2021, 99, 192–202.

26. Chen YH, Wang CC, Chen YY, Wang JH, Hung CH, Kuo YH. Low-dose nivolumab in advanced hepatocellular carcinoma. BMC Cancer. 2022;22:1153.

27. Meriggi F, Zaniboni A, Zaltieri A: Low-dose immunotherapy: Is it just an illusion? Biomedicines 2023 ; 11:1032, 2023

28. Trikha M, Sarkar L, Dhanawat A, Syed N, Gujarathi H, Vora M, et al. Performance of Low-Dose Immunotherapy and Standard-Dose Immunotherapy in Microsatellite Instability-High Metastatic Colorectal Cancer: Real-World Data (CLouD-High Study). JCO Glob Oncol. 2024 Aug;10:e2400141.

29. Guérin J, Nahid A, Tassy L, Deloger M, Bocquet F, Thézenas S, et al. Consore: A Powerful Federated Data Mining Tool Driving a French Research Network to Accelerate Cancer Research. Int J Environ Res Public Health. 2024; 21:189.

30. Heudel P, Crochet H, Durand T, Zrounba P, Blay JY. From data strategy to implementation to advance cancer research and cancer care: A French comprehensive cancer center experience. PLOS Digit Health. 2023 Dec 19;2(12):e0000415. doi: 10.1371/journal.pdig.

31. Heudel PE, Fervers B, Durand T, Chabaud S, Michallet AS, Gomez F, et al. Second primary cancers: a retrospective analysis of real world data using the enhanced medical research engine ConSoRe in a French comprehensive cancer center. Int J Clin Oncol. 2021; 26:1793–1804.

